# An unfortunate natural experiment in learning how to provide services to those in need: The case of Ukrainian war refugees with disabilities in Warsaw and Bucharest

**DOI:** 10.1101/2024.09.19.24314029

**Authors:** Monika Nowicka, Alexandra Deliu, Bogdan Voicu, Magdalena Szarota

## Abstract

When helping others, experience becomes important, especially in circumstances that involve interacting with a different culture—such as the ones implied in providing services to refugees. When disability is added to refuge, multiple types of experience become necessary, with the capacity for cross-sector collaboration being an asset. This paper explores the impact of the Russian invasion of Ukraine on the capacity of the Polish and Romanian organizations providing services to Ukrainian refugees in Warsaw and Bucharest, with a focus on disabled refugees. Based on 41 interviews with service providers/grass-roots organizations, it turns out that this unfortunate event served as a natural laboratory for practicing, acquiring, and increasing skills in multiple domains, leading to increased personal and institutional expertise. We inspect the differences between Warsaw and Bucharest, the first city having more experience in dealing with incoming flows of immigrants, the second being a newcomer in this respect. We also consider the distinction between public providers (public administration) and non-governmental organization entities, observing the upscaling of the latter. Implications for policy are considered within the framework of curtailing civic society under the illiberal wave.

## Introduction

The lessons learned by civil society and public service providers from dramatic events can significantly improve society in general and civil society in particular, by enhancing procedures, fostering cross-sectoral cooperation, and preemptively addressing crises (Alexander, 2022; Hashimoto, 2011; Sztompka, 2005). In this paper, we ask what lessons can be drawn from the *ad hoc*—or “spontaneous”—enthusiastic phases of helping Ukrainian refugees in Poland and Romania. We focus closely on the encounters of service providers with war refugees with disabilities and treat this negative life event as a natural experiment prompting societies and organizations for quick and efficient reactions.

In historical terms, the Russian invasion of Ukraine was probably not a surprise, since Russian armies have repeatedly occupied the area, both during the times of the tsars and in the soviet era. Earlier aggression in the Black Sea area (against Moldova and Georgia, for instance), as well as against Ukraine (such as the 2014 invasion of Crimea) preceded the beginning of the war in March 2022. However, the scale of the invasion was unprecedented in post-WW2 Europe, and the emotional response across the continent was considerable, affecting both citizens and decision makers, especially—but not exclusively—in the countries bordering Ukraine (Beauregard, 2022; Chudzicka-Czupała et al., 2023; Smith, 2021; Vus & Esterlis, 2022).

At the same time, despite people with disabilities in Western culture having high deservingness (Assouline & Gilad, 2022; Fang & Huber, 2020), particularly when disabilities are based on medical evidence (Geiger, 2021), post-communist European societies have experienced surges in miscontent from the disabled, about how they are treated by societies (Zdrodowska, 2023). The neglect of social policy costs in favor of economic growth and the existing barriers in accessing social services by the beneficiaries have prompted such reactions (Kiss, Primecz, & Toarniczky, 2022), in a context where special needs are still strongly stigmatized (Mikelsteins & Ryan, 2018) and little attention is paid to disability beyond the medical model (Šumskienė, Gevorgianienė, & Genienė, 2021). In times of recession, when state retraction was expected, the professionalization of non-governmental service providers was anticipated to prevail, thereby redressing the growing inequalities in accessing care (Dill, 2014), yet reports show that such organizations are still in their “infancy” phase (Popa, Vlase, & Morandau, 2016).

Sudden incoming flows of refugees typically put existing healthcare and welfare systems under stress, as demonstrated by other examples across Europe (Gottlieb & Schülle, 2021; Human Rights Watch, 2017; Schottland-Cox & Hartman, 2019), or are adverted in the early days of wars by healthcare specialists (Van Hemelrijck et al., 2022). Despite the international regulations concerning the helping of disabled refugees (Duell-Piening, 2018), the implementation is not always smooth, with the pitfalls becoming acute when hazards occur.

This creates a fruitful field for understanding how the sector of service providers has evolved in Poland and Romania—two EU member states that border Ukraine and are large enough to receive important flows of Ukrainians fleeing the war, while also sharing a common history of being communist and displaying anti-Russian sentiments.

Our paper is based on 41 interviews that were carried out in Warsaw and Bucharest in 2023, with service providers and regulators in the field of disabled refugees, including state agencies and non-governmental organizations (NGOs). We explore this corpus with the aim of understanding how such providers change over time, as well as seeing whether this unfortunate event has allowed them to increase their capabilities in terms of providing help to those in need, irrespective of whether they are refugees, immigrants, or members of the native population. We focus on upscaling and learning ways of doing; we explore whether and how they develop cooperation within their sector and beyond their sector; and we look for evidence that the acquired experience has been converted into newly acquired or improved competences. While other similar studies have stressed the general impact of the war on NGOs in the region (Bryan, Lea, & Hyánek, 2023; Petrescu, 2023), we go deeper, by considering cross-sectorial effects, intersectionality, and long-lasting changes, as well as comparing the public service providers in two capital cities. As negative societal events provide windows of opportunity (Bogdan, 2023; Hashimoto, 2011; Morgado, 2020), we expect increases not only in terms of internal practices and immediate know-how (Petrescu, 2023; Valentinov, Bolečeková, & Vaceková, 2017), but also in terms of cross-sectorial cooperation and the capacity to extend activity into other areas.

In the following, we depict a four-fold context, in which we concentrate on the local setups of organizations focused on migrants, while also considering service providers for disabled people, exploring questions of deservingness, and investigating the obstacles that NGOs face under the current illiberal wave. All contribute to the background of our story. A detailed presentation of our methodology explains our empirical sources, which are then extensively commented upon in the findings section. The concluding part of the paper discusses the dimensions of organizational change and upscaling that migration and disability NGOs and public service providers have experienced and explores how they signal organizational, policy, and research implications.

## Literature review

### Migration/refugee-centered organizations. Aims and approaches to disability

Both Romania and Poland have short histories of receiving migrants, as, historically, these countries have both been known for exporting migrants to other nations. In recent years, however, the number of (economic) immigrants in both countries has been growing (Górny, 2019; Leșcu, 2018).

In both countries, NGOs are the main providers of assistance and services to voluntary and non-voluntary migrants (Dragan, 2017; Roman, Manafi, Muresan, & Prada, 2023). These NGOs focus on activities that help refugees and migrants adapt to life in a new country, providing support in cultural, economic, and social areas, either complementing the actions of public institutions or replacing them when the state fails to act (Pawlak & Matusz-Protasiewicz, 2015). Although they are the main actors supporting immigrants in the receiving country, their activities are not limited to this area, since they are also focused on the broader society in the receiving country (Dragan, 2017; Nowak & Nowosielski, 2018).

The scope of NGO activities ranges from short-term help—such as emergency direct aid, legal assistance, and organizing various courses—to long-term involvement in advocacy and advising on policy development (Flanigan, 2022; Podgórska et al., 2023).

NGOs dealing with migrants and refugees only support people with disabilities relatively rarely, since migrants are typically able to work—and even when they are disabled, they are often invisible (Tofani, Berardi, Iorio, Galeoto, & Marceca, 2022). The intersection of migration and disability is most often experienced by people fleeing from at-risk areas, so NGOs most often encounter people with disabilities during refugee influxes. Even then, the level of support provided to refugees with disabilities depends on various factors, one key issue being the number of people in reception centers. As demonstrated by the example of the Greek islands, which are known for their struggle with significant influxes of people seeking international protection, a large number of refugees can render people with disabilities invisible (AMID, 2018). In Austria, where refugee numbers are significantly lower, there are established procedures and solutions for assisting refugees with disabilities (Kreinbucher, 2018). An additional challenge in providing help is the unclear legal status of these individuals, which can prevent access to many services (Kreinbucher, 2018). Other factors obstructing effective and efficient support include a lack of data and the absence of a standardized method for assessing vulnerability (Tofani et al., 2023).

### Disability-centered organizations. Aims and approaches to migration topics

Disability researchers have stressed the crucial role played by disability organizations in the post-socialist region, especially given the scarcity of state-provided services intended for disabled persons (Mladenov, 2017; Rasell & Iarskaia-Smirnova, 2011). Such disability organizations have therefore been patching significant gaps in the provision of essential medical, welfare, and societal support systems that have been neglected in the evolving capitalist economies of countries like Romania and Poland (Bungău, Țiț, Popa, Sabău, & Cioca, 2019; Kocejko, 2018). This need resulted from the state’s failure to offer such critical services, as well as the large costs that were subsequently imposed upon disabled individuals, rendering such services financially inaccessible in heavily privatized sectors.

This predicament disproportionately affected disabled people in post-socialist regions, who, for a variety of reasons, typically belong to the most economically disadvantaged segments of society. As a consequence, disability organizations have mainly focused their efforts on providing assistance along impairment-specific lines, rather than adopting a more inclusive cross-impairment approach (Mladenov, 2017; Popa et al., 2016; Rasell & Iarskaia-Smirnova, 2011). This tendency partly resulted from the structures of the available state funding grant schemes, which emphasized objectives such as facilitating employment for disabled individuals and integrating them into societal frameworks, primarily through work-related and integration programs. As a result, scholars in Disability Studies have noted the pragmatic approaches adopted by these organizations to secure resources for their vital activities, compensating for the inadequacies of the state. However, the emergence of the Convention on the Rights of Persons with Disabilities (CRPD) led to shifts in some disability organizations, both large and small, toward aligning their objectives with the values outlined in the CRPD. These organizations began advocating for the replacement of medical and charity models of disability with a more comprehensive human-rights-based model—a shift that aimed to address disability discrimination in a systematic way, acknowledging its intersectional and holistic nature (Baciu & Lazar, 2017; Kocejko, 2018).

Before the arrival of Ukrainian war refugees with disabilities (UWRwD) in Poland, Polish disabled-focused NGOs did not support migrants and refugees, which is reflected in the not very rich literature on health-related issues and migration and refuge. The researchers focused mainly on access to public health services and social security issues (Andrejuk 2017, Strzemieczna 2021).

The intersectionality between disability and migration appears only marginally in the literature on immigration relating to the two countries (Chłoń-Domińczak, 2020; Goga, 2019; Oltean & Găvruș, 2018; Sterniński, 2019) and, to the best of our belief, has not been thoroughly analyzed. The reports in the two countries depicting the provision of social services almost completely ignored questions of immigration prior to the current Ukrainian refugee crisis (e.g., Ministerul Muncii și Solidarității Sociale, 2023; Radulescu & Paulischin, 2017; Vereha et al., 2022), while the reports of recent years show a completely different side of the service providers involved in matters of dealing with disability and refuge at the same time (e.g., Centrone, Gromada, & Posylnyi, 2023; Petrescu, 2023).

### Experiences shaping the approaches toward aiding refugees

Civil society organizations provide services to those excluded by public policies and restrictions in the field of immigration (Ambrosini & Van der Leun, 2015), and their role in the integration of refugees and asylum seekers has been long studied (Bešić, Diedrich, & Aigner, 2021; Collini, 2022). In their activities, they must accord with the rules governing forced migration and the integration of displaced populations, which, at the same time, often result in existing policies or discriminatory practices being reinforced (Ambrosini & Van der Leun, 2015; Bird & Schmid, 2023).

In terms of their main activity, there are NGOs that specialize in service provision to refugees and asylum seekers and NGOs that mainly engage in advocacy. Similarly, depending on their members, there are majority-led and migrant-led organizations (De Jong & Ataç, 2017). Ultimately, NGOs are dominated by the need to balance state policies of exclusion with the need to offer a proper response to refugees, paying attention to basic human rights (Robinson, 2013). This dichotomy of care versus control is even more stringent when the (limited) funds available to NGOs are distributed/managed by public authorities, decreasing the propensity of NGOs to advocate or uphold stances against their (main) funders (Alejandro, 2006; Bendell, 2006; De Jong & Ataç, 2017; Lau, 2021).

The duality between perceiving refugees as victims/recipients of help, on the one hand, and as individuals with their own agency, on the other, seems to shape the attitudes of NGOs as well as their ways of engaging with their clients/beneficiaries, which are closely linked to the characteristics of the organizations. For example, (migrant-led) refugee community organizations contribute to the empowerment of refugees, while NGOs, as a result of their more formal ways of organizing and delivering services, instead tend to see refugees as recipients of help (Schnelzer, Franz, Mocca, & Kazepov, 2022). When refugees are seen as victims, this actually contributes to making them passive, by stripping them of their agency (Robinson, 2013).

Existing scholarship has drawn attention to some important elements, such as the attitudes and activities of local authorities (Mensink, Čemová, Ricciuti, & Bauer, 2018); the spaces of interaction between service providers and their clients/beneficiaries and the involvement of refugees and asylum seekers as an empowerment mechanism (De Jong & Ataç, 2017; Elmammeri, 2023); the communication, cooperation, and coordination between service provider entities (Schnelzer et al., 2022); the hurdles of bureaucracy and the need for adequate training for frontline social workers (Robinson, 2013); and the difficulties generated by working in hostile environments, where general attitudes toward immigration and forced migration are negative, especially when governments are not in favor of immigration and establish strict policies as a consequence (Vosyliūtė & Conte, 2019).

### The framework for curtailing civic society under the illiberal wave

In the 1990s and 2000s, Central and Eastern Europe were marked by a large deficit in civic participation, due to their totalitarian pasts (Fidrmuc & Gërxhani, 2008). Institutional steps toward encouraging the flourishing of voluntary associations were not always clear (Meyer, Moder, Neumayr, & Vandor, 2019). And while increases in participation have been reported in the more recent literature (Ekiert, Kubik, & Wenzel, 2017; Murray Svidroňová, 2020; Voicu, 2020), the steps forward have nonetheless been rather modest (Ekiert & Kubik, 2017; Foa & Ekiert, 2017), overlapping with at least a decade of rising populism, illiberal rule (Merkel & Lührmann, 2021; Szelenyi & Csillag, 2015), and aggression toward NGOs (Bryan et al., 2023; Hann, 2020; Pap, 2017; Piotrowski, 2020)

Paradoxically or not, at the same time, post-communist societies, including Poland and Romania, have started to recognize the important role of civic organizations in providing welfare services complementing those offered by the state, when needed, due to their greater flexibility and adaptability in terms of providing tailored programs (Dill, 2014; Mladenov, 2017; Popa et al., 2016; Rasell & Iarskaia-Smirnova, 2011).

As a result, there is now a complex setup, with two opposite trends: first, a tendency to expect the booming of the sector, due to its social role, especially given the stress accompanying the war and the emotional reactions towards Russian aggression; and second, a contrary backlash against the sector, due to the influence of illiberal ideology. Let us also note that the latter is coupled with a global perception of a decrease in deservingness relating to migrants, which preceded the Russian invasion of Ukraine (Blachnicka-Ciacek, Trąbka, Budginaite-Mackine, Parutis, & Pustulka, 2021; Kissová, 2017).

### Major negative societal events as structure of opportunity

Natural and human-driven disasters have been shown to sometimes provide windows of opportunity for better developing communities and societies (Arcaya, Raker, & Waters, 2020; Bănică, Kourtit, & Nijkamp, 2020; Hashimoto, 2011; Morgado, 2020). To cope with the traumatic consequences of disasters, including wars, societies need to react quickly in adapting existing tools to the new realities. Institutional adjustments and organizational development are likely outcomes. Following the social acceleration assumption, that societies always change in an accelerated manner (Bogdan, 2023; Rosa, 2009), we claim that war generates a structure of opportunities for further change, especially in terms of a more flexible civic society that is able to supplement a lack of state action—this, in turn, being hampered by bureaucratic procedures. Similar positions have been stated by other scholars, who see the unfortunate massive flows of refugees as part of the structures of opportunity (Özdora Akşak & Dimitrova, 2021), as possibilities for rectifying the public blame of illiberal discourse and as indicators of NGO resilience (Bryan et al., 2023).

## The organizational/institutional ecosystem in Poland and Romania prior to the war

### Migration

In both countries, the incoming flow of refugees was accompanied by a strong civic reaction, with regular citizens and NGOs helping them, especially at borders, and shaping the public authority response (Bogdan, 2023; Fomina & Pachocka, 2024). This was not unexpected, since, in at least some post-communist societies, NGOs have assumed the major role in terms of migrant integration over the past decade—something that reflects a lack of policy at the national and local levels (Bryan et al., 2023; Cech Valentová, 2018; Voicu, Deliu, Tomescu, & Neguț, 2018).

In **Poland,** the assistance and integration system for migrants relies on short-term decisions, due to the absence of a developed integration policy for the city. The ecosystem assisting migration involves various stakeholders, but the system is not entirely open and accessible to migrants in Poland Indeed, access is contingent upon the legal status of the migrant, with two main types of actors involved: city-run institutions offering access to social services and NGOs operating within grant financing systems (Wach & Pachocka, 2022; WCPR, 2024).

At the city level, the key institution involved in migrant integration is social welfare center, which provides support to migrants with refugee or subsidiary protection status, including Ukrainian war refugees, who have had temporary protection status since 2022. In Warsaw it is the Warsaw Family Support Center. Economic migrants with legal residence and access to the labor market are not covered, but are still able to seek support from Employment Office, another local government institution.

The crucial actors in this process are local and international NGOs, which offer various integration activities to migrants, including legal assistance, cultural integration, psychological support, professional qualifications, language courses, as well as social support for those facing financial challenges. An important and quite recent actor is the Konsorcjum Migracyjne/Migration Consortium, which is a democratic platform for NGO cooperation.

A number of NGOs in operation in Warsaw have been established and are managed by immigrants, the most significant being the Ukrainian House. It is noteworthy that an increasing number of NGOs are now employing migrants as part of their workforce (Ukrainian House, 2024).

In **Romania**, the main public institution in charge of controlling and supervising immigration is the General Inspectorate for Immigration—which is, in fact, a branch of the police force, under the rule of the Ministry of Internal Affairs (IGI, 2024). When it comes to the public provision of (social) services, the most important actor is the General Direction for Social Work and Child Protection. In Bucharest, the institution is subordinated to the Local Council. Another public provider of social services is the General Direction of Social Work of the City/Municipality of Bucharest, which is part of the city hall.

International organizations such as the United Nations High Commissioner for Refugees (UNHCR) or International Organization for Migration (IOM) are present in Romania and collaborate with local NGOs, developing programs for those arriving in Romania either as economic migrants or asylum seekers. On UNHCR’s website, the organizations providing support for immigrants are listed by location (Bucharest and other locations near the border). A significant stakeholder is the Coalition for the Rights of Migrants and Refugees, an entity specialized in advocacy, with NGOs as members. The number of NGOs in the field is therefore not as small as has been reported elsewhere (Flanigan, 2022, p. 10), and during the initial days of the conflict there was a sudden increase, due to the involvement of other civic organizations and NGOs that were devoted to providing social services (Flanigan, 2022). Indeed, civil society mobilized itself in order to contribute to the provision of solutions meeting the needs of the Ukrainian refugees coming to Romania. NGOs from various fields extended their services to refugees, thereby entering a learning and adaptation process, in which they had to overcome various barriers, such as those relating to administration and language (Petrescu et al., 2023). These types of barriers are not new, as they have already been reported by refugees and stakeholders alike, making service provision (for providers) and integration into the host society complex issues (Voicu et al., 2018).

As far as the authors are aware, there are no migrant-led grassroots organizations in Romania with visible involvement in advocacy and the aim of furthering discussions about the rights of migrants and refugees. This is explained at least in part by Romania being primarily a country of emigration, with scarce experience of hosting immigrants, whether they are economic migrants, asylum seekers, or refugees.

### Disability

Poland and Romania’s disability support systems exemplify the traits that are ingrained in post-socialist disability frameworks (Baciu & Lazar, 2017; Pamuła, Szarota, & Usiekniewicz, 2018). Scholars in Disability Studies have observed distinctive characteristics prevalent in this regional context (Mladenov, 2017; Rasell & Iarskaia-Smirnova, 2011). This approach portrays disabled people as culturally “other” and as economic burdens on society. The prevailing social policies are still mainly based on the medical and charity models of disability. The medical model views disabled individuals primarily as patients requiring medical intervention or rehabilitation, while the charitable model regards them as passive recipients of aid. Disabled people are valuable members of society and citizens of the state, but neither of these models depict them as such. Consequently, both models contribute to the segregation of disabled people into distinct impairment-related groups, thereby failing to identify and address their needs in an intersectional and comprehensive manner. This failure contradicts the requirements of the United Nations’ CRPD (2006), as ratified by Romania in 2010 and by Poland in 2012.

Persistent shortcomings in policy formulation and implementation—especially in education, healthcare, living conditions, and societal inclusion—perpetuate barriers that make it difficult for disabled people to lead independent and dignified lives (OHCHR, 2018). It is important to address these issues and ensure that policies and institutions are inclusive and supportive of people with disabilities. Alarming evidence points to nationwide instances where institutions aiming to support disabled people inadvertently contribute to their further isolation and, in some cases, subject them to violence (Bungău et al., 2019; McRae & Szarota, 2018). It can be argued that the responsibility for fulfilling the obligations outlined in the CRPD falls primarily on disability organizations in both countries, rather than state authorities (Bungău et al., 2019; Kubicki, Bakalarczyk, & Mackiewicz-Ziccardi, 2019). However, these organizations face significant challenges in taking action, due to limited funding and resources, relying heavily on grant schemes to exert influence. As a result, the provision of support for disabled people is predominantly concentrated in larger urban centers in Poland and Romania. In summary, despite variations in the impact of disability movements, both Poland and Romania are confronting systemic challenges that impede the establishment and maintenance of supportive ecosystems for individuals with disabilities, in alignment with the human rights and obligations specified in the CRPD.

## Hypotheses

Prior to the Ukrainian crisis, as a consequence of the flow of Syrian refugees, Valentinov et al. (2017) noted that this sudden challenge helped NGOs in Austria and Slovakia to define a clear identity and better structure their activities. A report on the changes in NGOs providing support to Ukrainian refugees focused on the Romanian NGOs financed by the French CARE foundation, concluding that such organizations report **upscaling** in terms of recruiting staff and investing in infrastructure, intensifying fundraising efforts, rethinking the structure of the organization, and providing more “tailor-made interventions” as a rule of organizational life (Petrescu, 2023). Other reports compare NGOs with the state, stressing the need for flexible interventions in the early days of conflicts (Bryan et al., 2023), something that requires increased speed (Bogdan, 2023).

We expect similar results and we go one stage further in inspecting the consequences, by looking for specific evidence of **intersectionality** and **cooperation** between sectors. We also consider the state of the sector prior to the war as definitory in terms of the observed impacts. When the war suddenly broke out, the efforts to provide for those in need were still in their initial phases in both countries. Adequate provision for both refugees and the disabled had yet to be achieved. Therefore, we expect the results for intersectionality to be even worse, with migration/refugee organizations and public service providers not adequately accommodating disabled refugees. We also hypothesize that the war resulted in important **changes to the everyday lives of service providers**, not only in numerical terms—with upscaling being expected, in order to face the flow—but also in terms of **deriving knowledge and good practices.**

Two years after the war started, with its ending still uncertain, the knowhow for improving existing methods likely exists “on the ground,” through trial and error as well as the ad hoc “good inventions” of various civil society and public actors, yet it nonetheless requires identification, labeling, and streamlining (e.g. through cross-sectoral recommendations). We also expect to observe an intensification of the cooperation across sectors; that is, organizations providing services to migrants and those dealing with the disabled are expected to engage in exchanges of knowledge and practices. In terms of the consequences for the disabled, we anticipate that social acceleration will have provided both the knowledge and tools for better service provision, including beyond situations of social emergency.

Finally, given that Polish society was already better prepared to react (Van Hemelrijck et al., 2022), we expect the processes of change depicted above to be found to a more sophisticated extent in Warsaw than in Bucharest.

## Data and methods

In our research, we applied two distinct methodological approaches: participatory research and rapid response research. Our emphasis was on defining local priorities and perspectives integral to the participatory approach, drawing insights from the work of Cornwall and Jewkes (1995). Simultaneously, the rapid response research approach aimed to swiftly develop responses to emerging problems, in alignment with the framework proposed by Stimson, Fitch, Rhodes, & Ball (2009). Ethical considerations, as highlighted by Van Brown (2020), and recommendations in Poland (Trzeszczyńska & Horolets, 2022), led us to focus on helpers, not on refugees. Purposive sampling guided our selection of stakeholders based on two criteria: experience in supporting migrants/refugees and experience in supporting people with disabilities. To maintain balance, we carefully controlled the representation of public and non-public actors.

The empirical data for this article was collected as part of the [anonymized project; we will add the exact information and link to the website after the reviewing process]. The fieldwork took place between July 15 and December 15, 2023, at two research sites, Warsaw and Bucharest. All the participants provided oral informed consent before the interview. Then the consent was repeated and recorded at the beginning of each interview. The research comprised three stages: preliminary research involving an initial workshop with stakeholders supporting either migrants/refugees or people with disabilities; individual in-depth interviews (41); and an extended group interview discussing hypothetical cases of refugees with disabilities. The fieldwork conducted at the two sites followed the same approach. Additional information was collected during research visits to service providers in Bucharest (3) and Warsaw (3).

The initial workshop served as a valuable resource for establishing connections, constructing a potential database of respondents, and gaining preliminary insights into the day-to-day challenges of professionals. This workshop facilitated the calibration of research instruments and adjustments to the list of potential respondents.

The most challenging aspect of the data collection proved to be the in-depth interviews with individual stakeholders. Factors such as busy schedules, fatigue, and overwork posed difficulties. In Romania, an additional factor was a lack of relevant cases, which hindered the experience of participants in dealing with such situations. In Poland, the delay was further exacerbated by Parliamentary elections, which caused some public officials to be reluctant to participate.

**Table 1.** Structure of the sample.

|  | Bucharest |  | Warsaw |  |
| --- | --- | --- | --- | --- |
|  | No. of<br>interviews | No. of<br>interviewees | No. of<br>interviews | No. of<br>interviewees |
| Representatives of local authorities<br>(providers of public services) | 6 | 8 | 2 | 2 |
| Representatives of NGOs in the field of<br>disabilities | 4 | 4 | 9 | 9 |
| Representatives of NGOs in the field of<br>refugees/immigration | 5 | 5 | 5 | 5 |
| Representatives of NGOs/stakeholders<br>extending their services to refugees from<br>Ukraine | 3 | 4 | 1 | 1 |
| Volunteers involved in offering help to<br>Ukrainian refugees | 2 | 2 | 1 | 1 |
| Representatives of public institutions<br>(above local authorities) | 0 | 0 | 3 | 3 |

Most interviews were one-on-one, with a few involving two or three respondents. The specificity of the research theme probably deterred participants, due to their limited experience with cases of refugees with disabilities, and the participation of multiple respondents in the same encounter was perceived as an attempt to maximize the gains of the researchers or help them in gathering more relevant information.

The response rate was relatively low at both research sites: 20 interviews were completed in Bucharest, after reaching out to 60 individuals, and 21 were completed in Warsaw, after approaching almost 50 potential respondents. Interviews were conducted face-to-face (6 in Romania, 9 in Poland), online (12 in Romania, 8 in Poland), or by phone (2 in Romania, 4 in Poland), according to the respondents’ preferences.

Following the completion of a minimum of 20 interviews, we organized an extended group interview with relevant stakeholders, moderated by an external expert. This workshop focused on discussing potential cases of refugees with disabilities as well as addressing issues such as available resources, access, and possible setbacks. The cases were defined by the researchers and provided to the participants and the moderator. This event served as a means of validating the research findings and enriching them with practical insights from local actors. Its added value resulted from bringing together stakeholders with diverse backgrounds— NGOs (in Poland and Romania), public administration (in Poland and Romania), and academia (in Romania)— as well as from fostering collaboration and reaching consensus on the given cases.

At each stage of the research, respondents were informed about the purpose and funding, and each participant provided consent for interviews and recordings. All interviews were anonymized to ensure confidentiality.

## Findings

### Strategies adopted by service providers

#### Warsaw

Both NGOs and public service providers were unprepared to help high numbers of refugees, including those with disabilities. The first stage of helping was based on chaotic responses and a high level of engagement, with mainly grassroots organizations taking part. The main frame of reference for the activities of public institutions was constituted by the legal provisions defining target groups and methods of assistance. When possible, public agencies and NGOs were inclusive in helping: citizenship was not an aid criterion for either, with both having programs for which participants were eligible as long as they legally resided in Poland. There was one exception, however: if a person wished to be eligible for state social security support due to having a disability, then a Polish disability certificate was necessary. In other instances, a declaration of disability was enough. The state agencies did not provide help directly; rather, they transferred money to other entities, such as NGOs or local governments, to directly organize help to UWRwD:

> *(Anonymized) directly does not provide this support in the sense of organizing accommodation, meals, or rehabilitation activities for individuals from Ukraine. Instead, (anonymized) outsources these services to other entities.* (PL19, M, agency of public administration)

Building networks of cooperation was a common strategy for local government agencies and NGOs. Organizations did not provide all the support needed: they usually focused on some aspects, and if a beneficiary required help beyond their scope, they knew where to redirect that beneficiary—for instance, one of the disability-focused organizations (PL06) cooperated with a migration organization when a translation was needed, with another organization providing food and another offering financial support, if needed. In turn, the organization supported others when they required equipment for people with physical disabilities:

> *Of course [we have allies], because they also work towards our goals, and that’s why very often such situations where we can recommend our Foundation to people, like recommending us from other foundations, we do the same.* (PL06, F, NGO focused on the disabled)

However, the cooperation between the non-governmental and public sectors was limited. The main focus was on applying for grants that were called for by public institutions. State institutions were perceived by NGOs as being only temporarily present and slow in organizing help—indeed, after some time, it was observed that they began to withdraw from the assistance (GI).

Local government, to some extent, and NGOs based their systems of helping Ukrainian war refugees with disabilities on the voluntary work of both Poles as well as Ukrainians. This was especially true during the first few months of helping, when the influx was high but there was a lack of human resources and experience as well as a lack of procedures:

> *we ask the refugees themselves who stay in the hostel to take care of such a person in these basic matters […] Sometimes we have friends who do this work […] but it’s not a systemic solution.* (PL04, M, NGO focused on immigrants)

Over time and with increased funding, the NGOs started to hire new employees and sometimes even open new offices.

It seems that the main differences in approach to helping Ukrainian war refugees with disabilities are not between public and non-governmental entities, but rather between those state entities that support refugees directly and those that support them indirectly. Direct contacts with beneficiaries created common experiences for actors helping refugees. However, these experiences were not converted into new procedures and regulations that could be used in the future, should Poland receive refugees again.

#### Bucharest

According to our interviewees, the story was similar in Romania with respect to the flow of refugees, with surprise, a lack of preparedness, and chaotic responses characterizing the first few days. Local NGOs, being more supple in terms of their organization, focused on migration issues, with those NGOs dealing with social services intervening and providing unstructured help. Even after the state agencies stepped in and organized refugee camps, the presence of NGOs—this time backed by their transnational branches—continued to be essential, especially in tailoring the interventions to the needs of refugees in general and UWRwD in particular. However, neither the NGO sector nor the state agencies had specific provisions for refugees with disabilities. Intersectionality was increasingly a hot issue, not only in relation to the need to recognize disability certificates, but also when realizing that many other facilities were actually missing from the service provision:

> *we are deficient in providing services to people with disabilities, of any kind of disability. People with physical disabilities are more visible, obviously. But we are still far from providing any services for people with disabilities.* (RO17, F, NGO focused on other issues than refugees or disabled)

Cross-domain cooperation (migration versus disability) was forced by the particular difficulties being encountered, mainly in terms of the capacity to support refugees. NGOs focused on immigration-related issues, including refuge, started to refer UWRwD to NGOs in the social sector more often, especially those dealing with disabilities. At least until mid-2023, the problem was not one of financing, but related to human resources, the burnout of service providers, skills, the availability of the proper tools and facilities, and the capacity for advocacy. The social acceleration assumption proved to be true, as one of our interviewees points out:

> *cooperation was due to overcrowding; that is the main dominant criterion. Access to funds was secondary because, in general, since the war started and until the end of this year, there have been more funds than the capacity to help […] At least that’s how I estimate it. That is, no one complained about… about a lack of… of funds. On the contrary, there were organizations that said, “oh, too many donors come to us”; there were organizations that had to give up some donors by directing them to others.* (RO14, M, NGO focused on migration issues)

Considering the public agencies, they developed creative solutions in order to deal with the lack of regulations as well as funding: bringing resources from home, improvising, and letting NGOs step in and provide the services in spaces run by the public agencies. Again, cooperation occurred mainly as a result of the need to provide services alongside overcrowding. Since this need was apparent early on, public providers were accordingly open to accepting help from NGOs from very early on.

Both state agencies and NGOs claimed that the legislation was ancillary, slow in evolving, and revealed little understanding of the actual needs of refugees (with disabilities). One of the interviewees clearly pointed out that:

> *If the Romanian state has decided to do this, to make this change, it did a good thing. In my opinion, it should have been done a long time ago because they got used to the idea that we have rights for too long. And suddenly, on May 1st, the state came with the ordinance […] you must integrate. In my opinion, this should have come gradually, it was a year to stay, you have the right, receive everything, and then […] they were pushed, as they say, onto the streets, alone. It was all of a sudden, that’s my opinion. They are still people who need help, they are still disappointed, disillusioned, frightened, worried. Yes, the change came too abruptly. I believe this change should have come after 3, 4, 5, 6 months.* (RO09, F, public refugee center)

To sum up, intersectionality was considered only in the later phases of refuge, when some cross-domain cooperation also started to exist. Nothing was programmatic—rather, it was reactive, resulting from the need to do something with and for those in need. NGOs came forward to help public service providers, and NGOs focused on migration were more open toward cooperating with the ones dealing with disability issues. We noticed little transfer of knowledge and good practices across domains, but we encountered accounts of cooperation within the domains—that is, service providers in a specific field mainly cooperated with other providers in that field.

### Changes observed among service providers

#### Warsaw

The first change for service providers was to have a new type of beneficiary:

> *Regarding war refugees with disabilities, how long has your organization been involved in this issue? For us, the moment of the outbreak of the war in Ukraine, the Russian invasion, was when we started thinking about such issues.* (PL15, F, NGO focused on disability)

This resulted in other changes and caused the NGOs to adopt strategies in response to the new demands. Disability and migration NGOs leveraged existing support networks to boost their efforts, forming new alliances to extend their capacities:

> *I saw a great need to be better prepared both in terms of expertise and, I mean, practically. I mean, I’m talking about the expertise that can be applied, right? […] I think it was a need for better information flow between organizations themselves. But we tried to fix it somehow by creating these shared files, whether on drives or creating this communication channel. We tried to somehow fix it and meet that need.* (PL15, F, NGO focused on disability)

Using networks provided opportunities to obtain knowledge and solutions:

> *And, honestly, I don’t know if we would have taken it on in such an organized way […] if it weren’t for the support from organizations that are more experienced in this field.* (PL15, F, NGO focused on disability)

Another aspect of change was expanding human resources. NGOs hired additional staff, many of whom were bilingual or multilingual and could overcome the language barriers between service providers and refugees:

> *I think here many organizations also experienced this sudden growth, and a lot of new people were hired, who often had to be trained because, as we mentioned, bilingual individuals were in high demand, and many of them, like me, completely changed their profession. So, it was necessary to quickly acquire a lot of knowledge.* (PL02, F, NGO focused on immigrants)

To address the varied needs of refugees, NGOs developed new services and projects, including bilingual hotlines, video services for the deaf, accessible information centers, and customized support services. This development required an expansion and strengthening of the administrative functions of NGOs, to manage operations effectively and ensure compliance with the various regulations:

> *Earlier, we never dreamed of such, such, such large amounts of money in the account, so to speak. Such large funding, it sounded nice. And that was such a leap, both financially and in terms of staff; our team expanded, and our experience as well. So, we had to learn a lot and gain a lot of experience. When I opened the point, which was again a new place, in a new location, something completely new, I approached it differently, also more calmly.* (PL07, F, NGO focused on disability)

Employees and volunteers gained new skills, particularly in handling sensitive and complex situations relating to the refugee crisis:

> *not only did we need knowledge and access to information to provide people with reliable, good, and up-to-date information, but also psychologically, it was challenging. We learned a lot in those first months, but we also understood that the knowledge and so-called hard skills we have need improvement. Also, as employees on the first line, we need improvement in the emotional sense to take care of ourselves, our rest, and our overall well-being. Improving qualifications in working with people in crisis is essential.* (PL02, F, Polish NGO focused on assisting Ukrainians)

A public service provider in Warsaw also experienced upscaling in human resources and the type and number of beneficiaries. However, the latter have decreased:

> *after February 24th, [anonymized] expanded by nearly a hundred employees due to the task assigned by the Voivode, involving Long-Term Stay Points for refugees from Ukraine. Initially, there were nine points, and currently, we have five, accommodating a total of approximately 1,020 individuals, give or take. (PL13, F, agency of public administration)*

The service providers that did not have direct contact with UWRwD created new programs and calls for grants for NGOS directly helping UWRwD, and also created funds, but did not experience upscaling in the human resources area.

Service providers experienced changes in the types and scopes of their activities as well as their numbers of employees. They gained a lot of knowledge and experience in helping refugees with disabilities. However, such change has been constant, due to the decreasing number of UWRwD and diminishing support from both public institutions as well as society.

#### Bucharest

Adaptability and upscaling were the bases for responding adequately to the needs of refugees from Ukraine, who constituted a new category of beneficiary. For NGOs in the field of immigration, the novelty resided in the country of origin, and for disability-focused NGOs, the status of the beneficiaries was new, with their multilayered vulnerability, rooted in being displaced and disabled at the same time. Both public and civil-society-based actors were involved in the provision of services to Ukrainian refugees with disabilities, and most often communication between the various entities proved to be key when dealing with the complex needs that emerged. The cooperation between public providers/structures and NGOs was an effective way of making sure that potential beneficiaries could access the services they required in time, due to the flexibility of the latter. Beneficiaries with disabilities were directed toward suitable service providers, but this was not straightforward and without hurdles:

> *The main difficulty is to find organizations that deal with disabilities, except for the parts of autism, where there are organizations that respond.* (RO12, F, NGO focused on other issues than refugees or disabled)

Cooperation was part of the learning process, especially for NGOs that were not in the field of immigration/refugees before the war, which had to adapt to the pace of change quickly. At first, the activities of NGOs were mainly directed by the needs they encountered directly in the field, but, as time went on, they gained structure and coherence as their knowledge about how to handle specific cases increased. Specific types of problems were encountered, and cooperation played a significant part in dealing with them, alongside constant communication with the beneficiaries:

> *With time, the organizations’ actions became more structured. We identified the needs and aligned ourselves more effectively. However, at the beginning, it was more of an improvisational effort […] We couldn’t predict the problems that would arise the following week […] In other words, after offering assistance, we need to get feedback to see if we did well or if we need to adjust something […] I thought that there were some lessons we learned from there, and we learned to see them. They are somehow knowledge, experiences that our international partners, who usually do this, brought to us.* (RO03, F, NGO focused on other issues than refugees or disabled)

In the case of public service providers, the considerable increase in the number of beneficiaries was usually not followed by an increase in the number of employees, which often resulted in overworking. In the case of NGOs, organizational growth was a direct consequence of expanding the pool of potential beneficiaries. This process was closely guided by the needs that were encountered “on the ground,” and, at least at first, there was a significant share of learning by doing. The existence of various funding schemes allowed for the upscaling process to take place. Teams were expanded with case workers, as well as with administrative personnel, to better pursue financing alternatives. Significant difficulties were associated with finding translators and, as many NGOs increased their scopes, it was sometimes hard to find the best-suited professionals, due to the high number of hiring offers. For public service providers, it was more a matter of being assigned to this new task associated with Ukrainian refugees, so situations that employees were not able to manage for long were not uncommon:

> *We came here for the first time, we got to know each other. In the meantime, we changed many people in the team because some didn’t want to, some couldn’t cope, some had various problems. We changed people; we weren’t a constant team.* (RO011, F, agency of public administration)

However, what started as exponential growth stopped over time, due to the diminishing of the available funds, leading to insecurities and uncertainties regarding the sustainability of some of the services that were provided and, accordingly, the sustainability of maintaining the newly acquired professionals. In this context, thorough documentation regarding case handling and good practices would probably have been an effective organizational tool for enhancing knowledge retention, beyond economic and systemic/structural fluctuations.

## Conclusion and discussion

Our research offers a cross-sectional description of experiences of helping refugees with disabilities in the face of negative events, requiring quick actions but, at the same time, creating structures of opportunity. The paper examines how and to what extent these structures of opportunity were used and also introduces the discourse of intersectionality into the field of supporting refugees with disabilities. Our research focuses on two metropolitan areas of countries with only limited experience of helping refugees, including those with disabilities. Furthermore, both these countries are post-communist countries, with relatively weak civil society spheres and ongoing struggles with the illiberal wave. The results of the research based on the two cases can be generalized to other countries with similar histories and experiences of (not) helping refugees with disabilities.

Based on our analysis, we argue that key institutional changes have occurred in the areas of: material and human resources, networks of cooperation, and programmatic upscaling. The upscaling was experienced by NGOs, and to a lesser extent by public agencies.

Regarding the material and human resource upscaling, the most significant factor related to the unprecedented financial support received by NGOs, especially those focused on migration and disability in Poland and those focused on migration in Romania. That support enabled the organizations to strengthen their human resources by hiring more qualified staff—both administrative personnel and case workers, including ones with a background in migration. The expansion allowed the NGOs to provide a broader range of assistance and acquire more physical space to accommodate more people in need. In Poland, some organizations were even able to open additional branches, while in Romania it was often the case that NGOs were present at the borders with Ukraine and the Republic of Moldova, to offer on-site assistance, which was a new type of experience.

In contrast, public providers did not experience the same level of upscaling, except for one in Warsaw. Due to their more structured nature, bureaucratic constraints, and generally less flexible approach, public services continued to operate within their original parameters. However, the new responsibilities assigned to them, such as distributing public funds to NGOs or organizing reception centers, broadened the scope of their material focus, even though their overall capacity did not expand as significantly as that of NGOs.

The organizations helping disabled refugees were prompted to extend their mission, especially through hands-on learning experiences. The organizations realized that they were very often not capable of addressing the multifaced needs resulting from displacement, disability, age, and other characteristics of refugees. As a result, we observed evolving collaboration between disability-focused and migration-focused NGOs in Poland and Romania. However, the nature of this collaboration varied between the two countries. In Poland, both migration-focused and disability-focused NGOs were strongly proactive in establishing networks and exchanging information. In Romania, by contrast, migration-focused organizations showed much more cross-sectorial initiative than disability-focused organizations. This difference between the two countries could be due to the comparative strengths of the disability advocacy sector, being stronger in Poland than in Romania.

The organizations acquired knowledge in terms of how to fundraise and how to manage large new projects. Due to the contact with a new kind of beneficiary, they gathered information about and built knowledge relating to supporting refugees with disabilities. In the case of UWRwD, neither legal residence nor access to many public services was an issue, but we observed that support providers had to learn how to help UWRwD in terms of accessibility and language issues, as well as approaches to defining, diagnosing, and treating various forms of disabilities. Gathering this knowledge was one of the most important dimensions of the upscaling of the organizations. However, most organizations have not managed to transform this accumulated practical knowledge into more established solutions, such as written procedures. This pattern confirms the existing trend within civil-society-sector organizations. Due to a lack of funds focused on long-term institutional development, these organizations usually do not have the capacity to turn their acquired knowhow into more organized approaches to action or systematic solutions. Such systematic solutions, as part of institutional knowledge acquisition, would be transferable across other organizations and would not depend on being retained solely by the people who produced that knowledge, meaning that they could stay with the organizations.

At the beginning of our research, we decided to distinguish between two categories of service providers— public ones and NGOs—and describe their experiences of supporting UWRwD. Our results revealed that, on the one hand, the public agencies were slower and not as flexible in helping refugees with disabilities. On the other hand, though, we could observe a difference between public agencies with direct and indirect contact with UWRwD. The former were mainly local government actors, with frontline workers having direct contact with UWRwD, making their experiences more similar to those of NGOs, since they were forced to introduce non-standard solutions while helping. Therefore, they were occasionally prone to contacting NGOs and building cross-sectional networks of support. They also experienced upscaling in terms of knowledge about supporting refugees with disabilities, which was a consequence of helping. As in case of the NGOs, however, this knowledge has not been codified.

Analysis of the collected data revealed that patterns of organizational growth can be observed both in the areas of human resources and programmatic assistance in both countries. The first stage of assistance, immediately after the outbreak of the war, was dominated by NGOs, which, despite limited resources and a lack of experience, started to support refugees with disabilities almost immediately. The consequences of these actions were overwork and employees burning out. The funds that quickly followed allowed for the employment of new employees, however, enabling the situation to be normalized and the activities to be expanded. Public actors subsequently joined the field of assistance, only after the creation of legal solutions allowing them to operate. Despite the presence of public actors, though, the main burden of assistance rested on the shoulders of NGOs.

In terms of programmatic assistance, in the first phase, the following pattern can be observed: recognition of the needs of refugees—diagnosis of one’s own capabilities—coming up with a solution—providing assistance or searching for an organization that can help and redirecting a beneficiary. This mode of operation contributed to the creation of a support network between NGOs, which furthered the provision in their areas of activity, allowing for more responsive and effective assistance. In the later stages, as the organizations became more experienced, the process of helping was smoother, as after diagnosing the needs of UWRwD, they usually knew where to direct a person. Recognizing the needs of subsequent immigrant cases allowed information to be collected about the best ways of helping refugees as well as coping with multicultural perceptions of disability.

This pattern is true for both Warsaw and Bucharest, but there were some specific differences between the two cities. The cross-domain cooperation was low, and it seems to have been higher in Bucharest than in Warsaw, while the within-domain cooperation was more common in Warsaw than in Bucharest. This was probably due to the fact that in Bucharest, the local authorities were more present in organizing help for UWRwD than in Warsaw, while the NGO sector supporting people with disabilities was more visible and involved in providing services to Ukrainian refugees with disabilities in Warsaw than in Bucharest.

Our research also has its limitations: we interviewed organizations in Warsaw and Bucharest, so our findings can be expanded to other large cities—but the experiences of helping at border areas, as well as in smaller towns, have not been reflected. Another limitation is the fact that the refugees with disabilities themselves and their caretakers who have daily routines of caring were not part of the research. Future research should be foremost devoted to gathering and analyzing the experiences of the refugees with disabilities themselves. Additionally, it should investigate whether and how cross-sectional cooperation continues to be implemented in the evolving national contexts of Poland and Romania, as well as over a broader geopolitical landscape. Another topic would be to examine how the discourse of intersectionality between refuge and disability is developing in Poland and Romania and other countries of Central and Eastern Europe.

In general, while the “unfortunate natural experiment” that we have documented resulted in capacity building, especially among the NGOs, and while it also led to previously unknown networking and cross-sectional cooperation between NGOs and public service providers, both in Poland and Romania, such upscaling trends could be unstable or reversible. This reversibility might be contingent not only upon the available funds, but also upon the human capacity to work in highly stressful conditions marked by constant change and uncertainty—even if the service providers and their staff are strongly motivated to help people.

As for policy recommendations, the key issues involves creating long-term policies for supporting refugees with disabilities and including refugees with disabilities in relevant laws and programs at both national and local levels.

## Data Availability

Data cannot be shared publicly because of conditions of the consent between researchers and the interviewees. Data are available from the Collegium Civitas Institutional Data Access / Ethics Committee (contact via) for researchers who meet the criteria for access to confidential data.

## Acknowledgements

This paper is funded through the Polish National Agency for Academic Exchange (NAWA) under the program “Urgency Grants Program”. Project title “Undisabling the refugee flow. Increasing the capacity of Polish and Romanian stakeholders to provide support to Ukrainian refugees with disabilities in the metropolitan areas of Warsaw and Bucharest.” BPN/GIN/2022/1/00102/U/00001.

